# A participatory approach to designing and implementing an occupational health intervention for the nail salon community in the Greater Philadelphia region

**DOI:** 10.1101/2023.06.10.23291234

**Authors:** Trân B Hùynh, Dương T Nguyễn, Nga Vũ, Lucy Robinson, Emily Trần, Nancy Nguyễn, Amy Carroll-Scott, Igor Burstyn

## Abstract

**Background:** The nail salon industry in the US comprises mostly immigrant-owned, small mom-and-pop salons that employ primarily first-generation immigrant workers from Asia. Because of the cultural and language barriers, both owners and workers may not avail themselves of the occupational safety resources. We formed an academic-community partnership to co-design a feasibility study and multi-level occupational health intervention for Vietnamese-speaking salon owners, workers, and community-based organization.

**Methods:** The intervention for each salon included 1) two-hour in-person training covering chemical safety, infection control, musculoskeletal prevention, and workers’ rights for both the owners and their employees, 2) a tailored recommendation report for the owner, and 3) check-ins with the owners during the three-month follow-up. Community partner was trained to deliver the in-language training with technical assistance from the research team. Baseline and post-intervention individual data about health symptoms and behaviors, as well as personal chemical exposures were collected and analyzed.

**Results:** A total of 44 participants from 12 consented salons enrolled in the study. One salon dropped out follow-up due to change of ownership. Analysis of the differences between post-and pre-intervention showed a tendency toward reduction in self-reported symptoms in the respiratory system, skin, and eyes, neurotoxicity score, as well as some chemical exposures. We could not rule out seasonality as an explanation for these trends. Increase in self-efficacy in some areas was observed post-intervention.

**Conclusions:** Our study demonstrated a successful academic-community partnership to engage community members in the intervention study. While the intervention effects from feasibility study should be interpreted with caution, our preliminary results indicated that our community-based intervention is a promising approach to reduce work-related exposures among nail salon worker workers from Asia.

## INTRODUCTION

Nail salon workers (NSW) are members of virtually every community in the United States (U.S.) and experience a constellation of occupational safety and health hazards that are not adequately addressed. Estimates of the number of NSWs ranged from approximately 126,000 (government source) to 400,000 (industry estimate) (NAILS Magazine, 2018; Sharma et al., 2018). This wide range reflects the challenges of capturing accurate employment data among this largely immigrant and refugee workforce. Approximately 80% of NSW are women and immigrants from Asia, with more than half of Vietnamese descent (Sharma et al., 2018). The chief occupational health and safety concerns among them include chronic exposure to chemicals, notably volatile organic compounds (VOC) such as ethyl acetate, n-butyl acetate, toluene, benzene and methyl methacrylate (Quiros-Alcala et al., 2019; Zhong et al., 2019). Phthalates and organophosphate esters (OPEs) were also detected in NSW’s breathing zone (Craig et al., 2019; Nguyen et al., 2022). These exposures have been linked with respiratory and dermal irritations, headaches, cognitive impairment, adverse endocrine and reproductive health (Ma et al., 2019; Nguyen et al., 2022; Quach et al., 2008, 2015, Quiros-Alcala et al., 2019). Other concerns include ergonomic hazards and exposure to blood-borne and airborne pathogens.

Despite calls to reduce workplace exposures among NSWs, there have been few documented intervention studies (Garcia et al., 2015; Roelofs C et al., 2010; Ward et al., 2022). Among them, two (a pilot study and a full randomized intervention study by the same research team in California) were culturally tailored to Vietnamese NSWs and owners, focusing on chemical safety (Quach et al., 2018a, 2018b). The randomized intervention study by Quach et al. (2018) utilized the train-the-trainer approach, in which the research team trained the owners in a day-long workshop in Vietnamese and the owners then trained their employees. The intervention was reported to improve chemical safety knowledge and behaviors. The study did not detect differences in the measured chemicals (toluene, methyl methacrylate, and total volatile organic compounds). Another intervention by Lux et al. (2014) was implemented by a health department and focused on infection control practices. It included multilingual educational materials, reminders sent to salons, outreach visits, and an inspection. Lux et al. (2014) reported a reduction in inspection infractions in both the intervention and control groups. However, a larger decrease was observed in the intervention compared to the control group. This effect may be due to the distribution of the educational intervention materials (Lux et al., 2014). Lastly, Meyer et al. (2015) developed training for cosmetology schools and their trainees. The materials included chemical exposure, infectious diseases, and other physical hazards and their control measures. The authors reported an increase in the knowledge assessment between post- and pre-tests. A positive impact on behavior such as use of personal protective equipment (PPE) was also noted. Overall, educational interventions tend to improve knowledge and self-reported safety behaviors. Improvements of the salon environment, such as product substitution or ventilation upgrades, were not assessed in the literature.

On the East coast, particularly the Greater Philadelphia region, several needs assessment studies were conducted with the nail salon community (Freeland et al., 2021; Huynh et al., 2019; Ma et al., 2019). We are a partnership of academics and a local grassroots organization, Vietlead, who co-developed and piloted a healthy nail salon intervention in the Philadelphia area. While the main purpose of this paper is to present our community participatory approach, feasibility measures and lessons learned to inform the larger study, we will also report effect estimates based on our behavioral survey, evaluation of respiratory and neurological symptoms, and personal air quality monitoring (which can help plan adequately powered future work).

## METHODS

### Study overview

Figure 1 illustrates the overview of our study’s workflow and design. The design is a one-arm trial where the intervention was delivered to everyone. The primary outcomes were feasibility measures, such as recruitment and retention rates and assessing the logistical challenges to inform a larger study. Secondary outcomes were individual health and behavior, observation of working environment within the salons, and monitoring of personal exposures. Pre-(baseline) and post-intervention data were analyzed to estimate the potential intervention effect, while noting that this pilot work was not powered to test the effectiveness of the intervention and the absence of a control arm, leaving us with the aim of estimating potential effect size and variance for the purpose of design of future evaluation of the interventions. We also conducted parallel qualitative process evaluation to better understand the perceived benefits and barriers to implementing the recommended practices (Huỳnh et al., 2023). Each participant was compensated $50 each time they completed the survey, $100 if they wore the air monitors, and $100 for attending the training.

**Figure 1:**
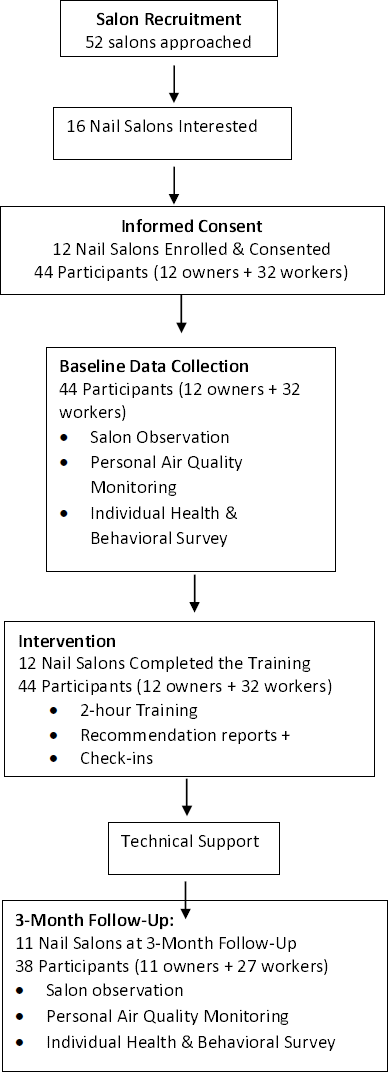
Schematic of the study

The ethics of our protocol was reviewed and approved by Drexel University Institutional Review Board (IRB) (Protocol # 2104008506).

### Community partnerships

We developed and implemented this study in partnership with Vietlead, a local community-based organization serving the Vietnamese community in the Philadelphia and South Jersey areas. Prior to our collaboration on this study, the academic team and Vietlead already had worked together on a community needs assessment (Freeland et al., 2021) and the initial testing of the training content (Huỳnh et al., 2023). Our approach is grounded in community-based participatory research (CBPR) (Wallerstein et al., (editors), 2018) where the academic team and Vietlead were involved in all steps of this study including grant proposal and study protocol development, creation of the intervention and survey tools, recruitment, intervention delivery and outcome evaluation. Team members who speak Vietnamese (the PI and the project manager) met weekly with Vietlead staff to discuss project’s progress and decision-making. The meetings were conducted in Vietnamese and English so our Vietnamese collaborators could feel comfortable providing feedback. This reflected our community-centered collaboration and ensured that community voices were heard and carried weight (Thompson et al., 2021). The PI met with collaborators who do not speak Vietnamese separately.

### Healthy Nail Salon Intervention and Delivery

We developed the intervention using two frameworks. The first framework is the hierarchy of control -- a commonly used approach to minimize exposure to workplace hazards in occupational health and safety field -- that consists of five tiers in order from the most to the least effective: eliminate the hazards entirely, substitute them with safer alternatives, install engineering controls to reduce exposure level, implement administrative controls, such as training, and, lastly, the use personal protective equipment (PPE) (NIOSH, 2023). While the hierarchy appears to be a linear approach, effective control of hazards in a dynamic workplace often requires a combination of measures from different tiers. Our second theoretical foundation was the Social Cognitive Theory (SCT) which posits that individual behaviors are influenced by their environment (Bandura, 1988). The SCT is often represented by a triangle describes the dynamic and reciprocal interplay between individual characteristics (i.e., knowledge, attitudes), behavioral factors (i.e., skills, practice, self-efficacy) and the environmental factors (i.e., social norms, access to resources) (Glanz et al., 2008). Our prior research with the nail salon community has pointed us to lack of knowledge of hazards and safe practices among NSWs and salon owners (Freeland et al., 2021; Huynh et al., 2019). Additionally, the salon management practices and lack of resources were identified as some of the institutional challenges (Huỳnh et al., 2023; Huynh et al., 2019). We developed training materials based on nail salon safety materials from government sources (namely, OSHA, EPA, and California Healthy Nail Salon Collaborative). We collaborated with a Vietnamese production company in California (3888 Films) to produce three short culturally tailored videos (about three minutes each) on infectious disease prevention, chemical safety, and musculoskeletal disorder prevention in Vietnamese with English subtitles. We also produced two bilingual animated movies on worker rights in collaboration with the Studios of New Mexico State University Innovative Media Research and Extension. The entire two-hour training covered four main topics. Each topic started with an overview video and more details for each topic. The videos are available on our bilingual website (healthysalonsproject.org).

The PI (TBH), project manager (DTN), and Vietlead’s trainer (NV) co-developed the power point slides training. It should be noted that all three researchers (TBH, DTN, NV) have lived experiences of the nail salon environment either as former nail salon workers, or growing up with family members who are nail salon workers. The training was initially drafted in English and then translated into Vietnamese. The academic team worked closely with Vietlead trainer to refine the Vietnamese content based on her feedback and then translated the final version back to English for participants more comfortable receiving the training in English, and to ensure that the meaning was not altered in translation. We tested the training materials with a small group of Vietnamese nail salon workers and owners online (because of the COVID-19 pandemic). Vietlead trainer also practiced the training multiple times at the office and a nail salon. Vietlead trainers delivered in-person training at the salons. We also conducted synchronous online training to a small group of participants (primarily young English-speaking participants) who were not able to attend the in-person training due to scheduling conflicts.

Another component of the intervention was the salon environment assessment checklist, which was designed for the owners to assess their salon’s environment. The academic team initially developed checklists to help owners identify areas for improvement. However, after testing the tools with one owner, we learned that the tool was too complicated for a person without prior training in occupational health and safety. Our community trainer consequently suggested that the academic team conduct the salon observation and produce a recommendation report for owners. Lastly, we added follow-up consultations with the owners to go over the recommendations and monthly check-ins during the 3-month follow-up. In summary, our intervention for each salon consisted of a two-hour training, a recommendation report based on salon environment observation, one consultation to go over the report, and two follow-up check-ins with the owners.

### Recruitment

We relied heavily on Vietlead’s extensive connections within the community to recruitment Vietnamese salons in the Philadelphia area. We also posted flyers near Vietlead’s office located inside a strip mall that houses other Asian small businesses (i.e., restaurants, grocery store, hair salon). We also paid for advertisements in a regional Vietnamese language magazine, visited salons near Vietlead’s office, and made cold calls. Word-of-mouth among Vietlead’s staff and volunteers, clients and their family members happened concurrently. Recruitment started in July 2021 and lasted through November 2021.

### Eligibility & Informed Consent

We recruited Vietnamese nail salons in the Philadelphia area within the state of Pennsylvania. It was important for us that the owners agreed to participate in the study to ensure their collaboration and minimize the threat of retaliation against workers due to participation in our research. We asked the owners for permission to recruit their employees, typically over the phone.

If emails were given to us, we would send a copy of the informed consent in advance. We went over the informed consent process at the salons. Participants were provided with a hard copy of the consent form. We developed a short presentation in Vietnamese (about 10-15 minutes) highlighting important points from the consent form. We made sure there was enough time to answer questions from the participants. Vietlead staff were trained to conduct the informed consent process and shadowed the academic team to the first few salons. Then they did most of the recruitment and consenting on their own. With English-speaking employees, the academic team went over the consent form with them in English. Written consent was obtained from each participant.

### Health and behavioral survey

The survey consists of three main sections: health symptoms, current practices, and demographics. We used validated surveys if available and developed our own questions otherwise. In the heath section, we asked about their general health, respiratory symptoms using the European Community Respiratory Health Survey (ECRHS) short screening questionnaire (ECRHS, 2018), skin symptoms adapted from the Nordic Occupational Skin Questionnaire (NOSQ) (Shamout & Adisesh, 2016), eye symptoms, musculoskeletal discomfort from the Nordic Musculoskeletal Questionnaire (Kuorinka et al., 1987). We used the 34-question version of the Euroquest questionnaire to screen for neurotoxic symptoms, although it should be emphasized that the use of this screening tool should not replace clinical diagnosis (Carter et al., 2002; Karlson et al., 2000; Kaukiainen et al., 2004). Responses were captured as never (0), sometimes (1), often (2), and very often (3). The total can range from 0 – 102 or analyzed by domains (neurological symptoms, psychosomatic symptoms, mood symptoms, memory and concentration, tiredness, sleep disturbances). Each domain score is calculated by the sum of score divided by the number of questions in each domain (Kaukiainen et al., 2004). The behavioral section includes questions about current practices (categorical) and self-efficacy (continuous). The demographic section includes smoking and drinking habits. Copy of the questionnaire is included as Supplemental Material 1.

The health, behavioral and demographic data were collected through a 30 to 45-min phone survey at baseline and 3 months after the intervention. The survey was conducted in Vietnamese (n=40) and in English (n=4) by trained bilingual research assistants from both the academic team and Vietlead staff. Study data were collected and managed using REDCap electronic data capture tools hosted at Drexel University (Harris et al., 2009, 2019).

### Environmental exposure

In each nail salon, personal air quality data were collected for two workers on two workdays at both baseline and 3-month follow-up (a total of up to four measurements per person). The days were selected to reflect one busy day (a Friday, a Saturday, or a Sunday) and a less busy weekday, at both baseline and follow-up. We asked the employees to wear a passive air-sampling monitor (546 Organic Vapor Badge, Assay Technology) near their personal breathing zones. On each sampling morning, a team member visited the salon to set up the equipment when the salon was open or at the beginning of the participating employee’s shift. We then came back to retrieve the air monitors either near the closing hour in the evening, or when the employee’s shift ended. Sometimes the salons may open longer than the employee’s shift. We also instructed the participants to fill out a form to record services they did for that day (Supplemental Material 2). We sent the badges to Assay Technology (an American Industrial Hygiene Association (AIHA) accredited lab) for analysis for acetone, ethyl acetate, methyl methacrylate (MMA), and toluene. These chemicals were selected based on previous nail salon studies (for comparison purpose) and their known toxicity and budgetary constraint. On average, a field blank was used for each salon per campaign. The badges were brought back to the lab for processing (i.e., checking samples and filling out the chain of custody forms) and were shipped within the timeframe indicated by Assay Technology.

### Statistical analysis

Summary measures for the individual health and behavioral survey items, knowledge and self-efficacy scores, neurological outcomes, and personal air quality measurements were computed for the baseline and follow-up periods. We estimated the differences between the two periods using conditional mixed effects models to adjust for correlation within salons and across time for individual participants. For binary heath outcome measurements, relative risks were estimated using a modified Poisson regression model (Zou, 2004). Current practices were grouped into “usually or always” versus “never or seldom or about half of the time” to create a binary outcome. For continuous self-efficacy measures, we used linear regressions, and reported mean differences. For neurological measures and personal air quality measures, we analyzed the outcomes on log scale based on the observed empirical distributions and lack of apparent normality. Personal chemical exposures below the laboratory limit of detection (LOD) were substituted with LOD/2 (Hornung & Reed, 1990). For the log-linear models, we reported the estimated multiplicative changes in the average from baseline to follow-up.

## RESULTS

### Recruitment & Loss to Follow-Up

We reached out to 52 nail salons, 16 of which expressed interests in participating. Four salons did not meet inclusion criteria due to being located in either New Jersey or Delaware, leaving 12 nail salons to enroll, our target number. The recruitment took us close to six months. Only about a fifth of contacted salons were eligible and recruited. A total of 44 participants (12 owners and 32 employees) consented to participate in the study. It is worth noting that the numbers of contacted salons and nail salon workers reached may have been greater than noted above, since our community partner reported having many informal conversations with community members about our project, while interacting with them regarding other matters. Most frequently reported reasons for refusal included inconvenience, lack of time, and salons perceived as being too old to upgrade. In some instances, the workers wanted to participate in the study but said that their owners did not. Sometimes, some but not all co-owners (from the same family) wanted to participate, due to fear of getting involved in the research; in such cases, the salon was not enrolled.

We completed baseline data collection and delivered the in-person training to all salons. One salon dropped out of the study after the training due to change of ownership; 11 salons completed the entire program (training and consultation) and post-program data collection. Four employees in the dropped-out salon naturally did not participate in the follow-up interviews.

### Environmental, behavioral, and health outcomes

Table 1 presents demographic characteristics of the participants for 11 completed salons (N=38). A full demographic table for all participants (12 salons, N=44) is included in the Supplemental Materials. Most participants were women (82%), born outside the US (90%), had either high school or lower education (79%), and reported difficulty speaking English (58%). The median age was 43 years; the median years in the US was 17 years. The median year in the nail industry was 11, median number of days worked in a week was 6 and hours worked per day -- 9. Just over a fourth of the participants reported not having health insurance and have not visited a doctor in the last 12 months.

**Table 1:**
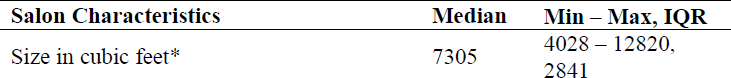
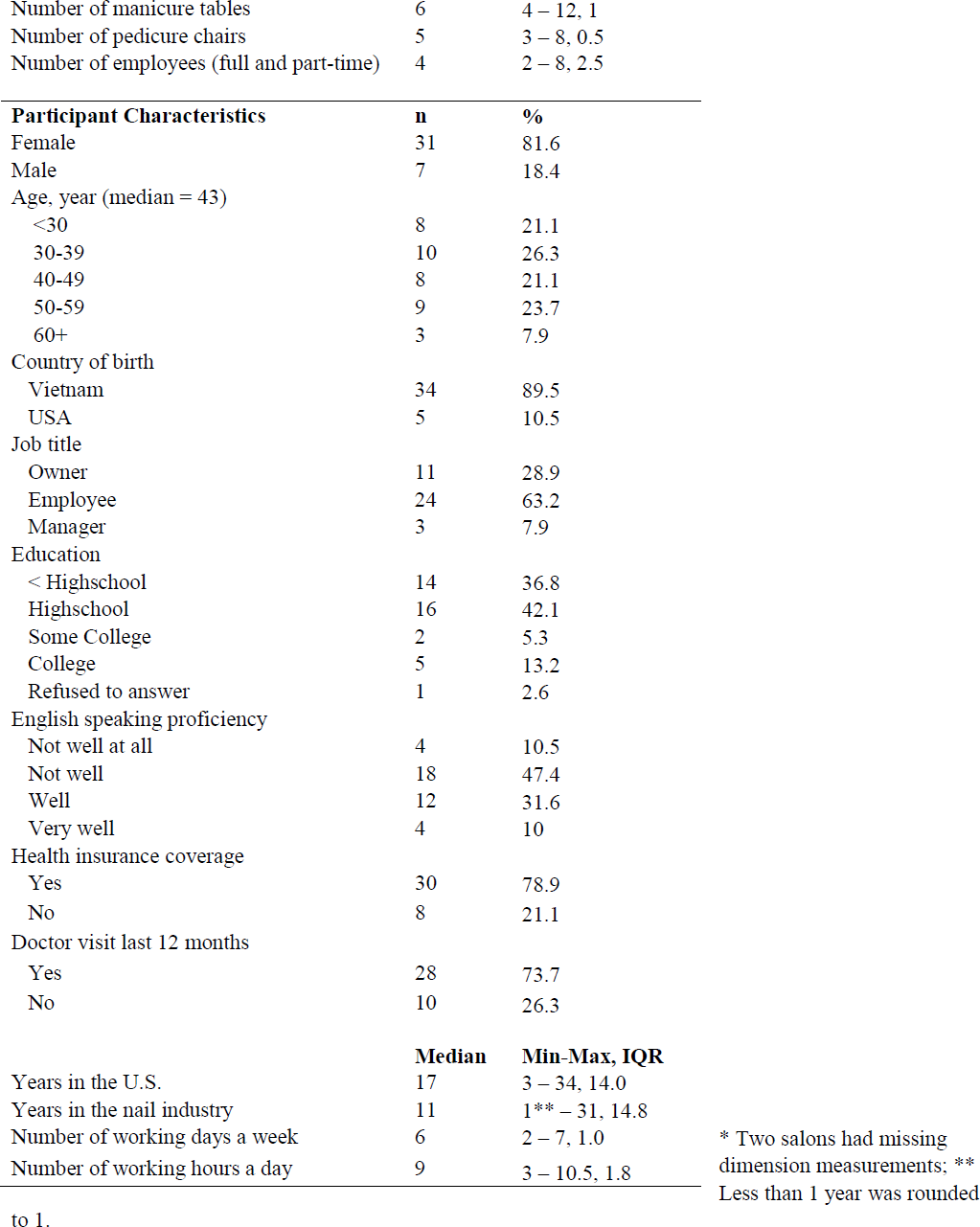
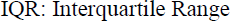
Salon characteristics and participant demographics from 11 enrolled salons in Philadelphia area (N=38) (One salon dropped out of the study)

Table 2 reports on self-reported symptoms related to the respiratory system, skin and eyes, and musculoskeletal disorders. At baseline, the most common respiratory symptoms reported were hay fever (48%), cough when sleeping (18%), and shortness of breath after strenuous activity (21%). Some participants reported that respiratory symptoms improved when away from work, most notably chest tightness (8%) and hay fever (16%). Approximately 40% of participants reported having eczema, most commonly at the hands (18%). A third reported eye irritation. About half of the participants who reported having eczema and eye irritation noted that their symptoms improved when away from work. Analysis of the relative risks (RR) between the post-vs. pre-intervention showed a tendency towards reduction for most respiratory symptoms: aRR=0.93, 95% CI: 0.51, 1.69; aRR=0.50, 95% CI: 0.12, 2.02, for chest tightness. The adjusted RR for eczema showed reduction (0.77, 95%CI: 0.39, 1.53) but symptoms for hand eczema and eye irritation appeared to increase after intervention (aRR=1.94, 95%CI: 0.99, 3.80, aRR=1.34, 95% CI: 0.68, 2.66, respectively). About a fifth of skin and eyes symptoms were reported to improve when away from work! The intervention did not appear to impact the reported number of symptoms that improved when away from work. For musculoskeletal symptoms (Table 2), we observed reduction of symptoms and adjusted RR after the intervention for most symptoms (e.g., neck pain, pain in elbows, hands, upper and lower back), except hip pain and pain in one or both shoulders during the last 12 months. Most participants reported an increase in pain that prevented them from doing normal activities for most areas of the body asked.

**Table 2:**
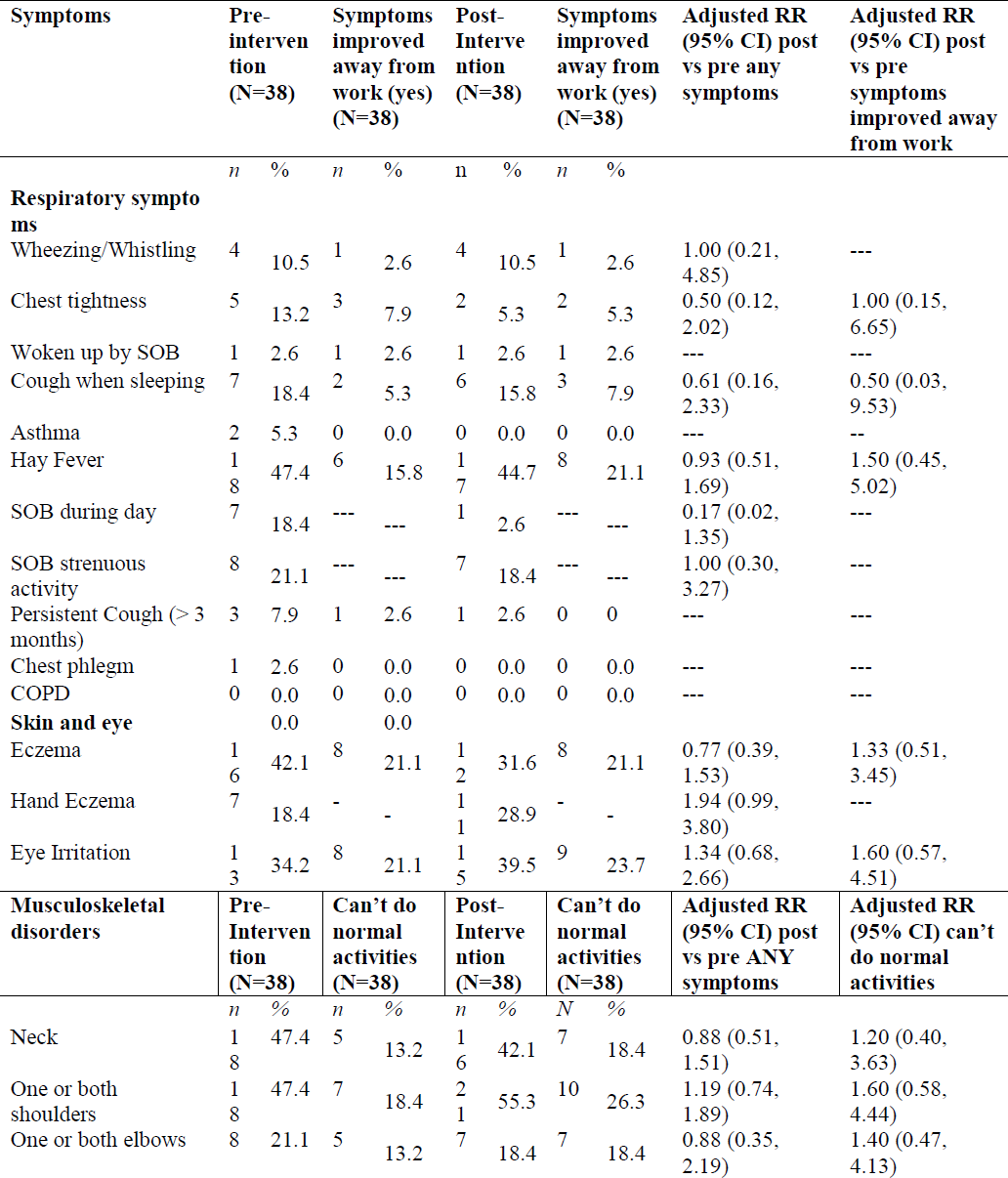
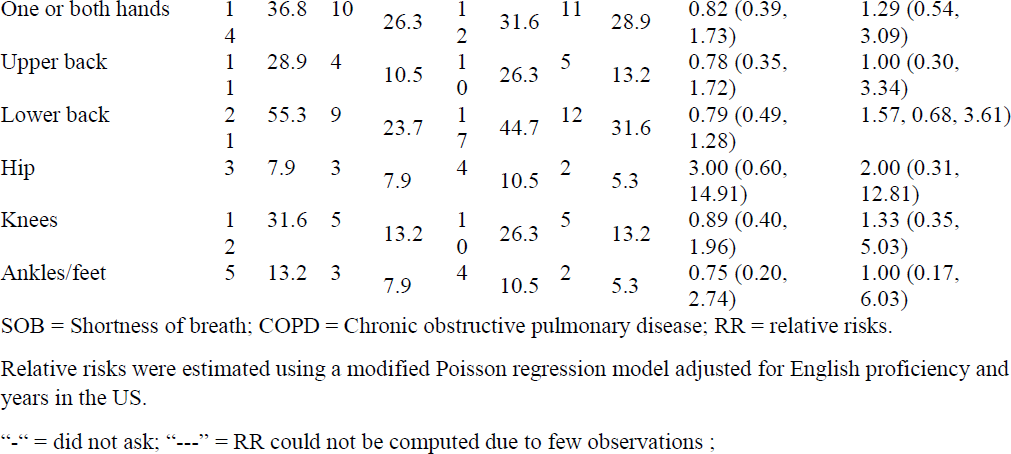
Prevalence of self-reported respiratory, musculoskeletal, skin and eye symptoms. The total of participants with completed baseline and follow-up data is 38.

We noted a reduction in the neurotoxicity scores in all domains, albeit with small differences (Table 3). For example, largest reduction was observed for memory domain at 6% (0.94; CI: 95%CI: 0.81, 1.09).

**Table 3:** Summary statistics and mean difference of the neurotoxicity score based on the 34-item Euroquest questionnaire.

| Domains | Pre-intervention |  | Post-Intervention |  | Mean ratio (95% CI) |
| --- | --- | --- | --- | --- | --- |
|  | Mean | SD | Mean | SD |  |
| Neurological | 0.30 | 0.28 | 0.29 | 0.28 | 0.99 (0.90, 1.10) |
| Psychosomatic | 0.38 | 0.32 | 0.33 | 0.31 | 0.97 (0.89, 1.07) |
| Mood | 0.37 | 0.42 | 0.34 | 0.32 | 1.00 (0.88, 1.12) |
| Memory | 0.66 | 0.55 | 0.56 | 0.47 | 0.94 (0.81, 1.09) |
| Tiredness | 0.64 | 0.74 | 0.62 | 0.76 | 0.97 (0.80, 1.19) |
| Sleep Disturbance | 0.53 | 0.47 | 0.54 | 0.52 | 1.00 (0.87, 1.15) |
| All questions | 0.45 | 0.33 | 0.41 | 0.32 | 0.98 (0.88, 1.09) |

Note: Missing values were imputed using row means.
SD = standard deviation; Outcome was analyzed on log-scale in the linear models that calculated within-person changes, akin to paired t-test; adjusted for English proficiency and years in the US.

We observed increased frequency of self-reported gloves use during services but a reduction use mask use, stretches and going outside to get fresh out post-intervention (Table 4). Self-efficacy increased in all assessed areas, except self-efficacy to clean foot spa (Table 4). The largest improvement in self-reported self-efficacy was with respect to protecting yourself from chemicals mean difference of 8.2 points, (95%CI: 1.5, 14.9), followed closely by adjusting body position to prevent pain 7.8 points (95% CI: 0.7, 14.9).

**Table 4:** Change in current practices and self-efficacy after follow-up.

| Current practices | Pre-intervention |  | Post-Intervention |  | RR (95% CI)<br>post vs pre* |
| --- | --- | --- | --- | --- | --- |
|  | n | % | n | % |  |
| Frequency of glove use for manicures in a typical day (usually/always) | 13 | 34 | 14 | 37 | 0.92 (0.46, 1.81) |
| Frequency of glove use for pedicures in a typical day (usually/always) | 32 | 89 | 35 | 95 | 1.04 (0.93, 1.16) |
| Frequency of mask use in a typical day (usually/always) | 38 | 100 | 37 | 97 | 0.97 (0.91, 1.03) |
| Frequency of performing stretches in a week (daily/4-6X per week) | 19 | 50 | 22 | 58 | 1.20 (0.73, 1.98) |
| Frequency of going outside to get fresh air in a week (daily/4-6X per week) | 19 | 50 | 22 | 58 | 1.82 (1.04, 3.17) |
| Self-efficacy | Pre-Intervention |  | Post-Intervention |  | Mean difference |
|  | Mean | SD | Mean | SD |  |
| Confidence in knowledge of cleaning and disinfection as required by State Board | 75.32 | 21.95 | 83.21 | 14.63 | 6.30 (-1.83, 14.42) |
| Confidence in ability to clean reusable tools | 83.62 | 14.81 | 87.62 | 10.64 | 5.16 (-1.07, 11.38) |
| Confidence in ability to clean foot spa | 86.19 | 14.63 | 87.68 | 12.38 | 0.72 (-6.05, 7.49) |
| Confidence in ability to safely handle blood | 84.49 | 15.64 | 88.35 | 11.14 | 4.63 (-1.99, 11.24) |
| Confidence in ability to adjust body position to prevent pain | 76.03 | 19.93 | 82.95 | 12.26 | 7.78 (0.67, 14.89) |
| Confidence in ability to keep yourself safe from chemicals at work | 75.11 | 16.78 | 83.26 | 11.76 | 8.17 (1.49, 14.85) |
| Able to talk to someone about work-related health concerns | 80.70 | 15.30 | 80.53 | 17.21 | 2.30 (-4.47, 9.07) |

Note: missing values were removed using na.rm=T
Relative risks were estimated using a modified Poisson regression model adjusted for English proficiency and years in the US.

Table 5 suggests that there may have been a reduction in some chemical exposures between the baseline and the follow-up evaluations. The most convincing evidence of reduction in personal exposure was for MMA, which dropped by 25% (ratio=0.76, 95% CI: 0.47, 1.23), personal exposure to acetone reduced on average by about 25% in the follow-up period (ratio=0.78, 95% CI: −50%, 9.1%) and ethyl acetate 6% lower (ratio=0.94; 95%CI: 0.68, 1.30), but the effect estimates were not precise. All toluene measurements were below the laboratory detection limits of between 0.11 – 0.20 ppm.

**Table 5:** Change in personal exposure to volatile chemicals (in ppm, where appropriate) for 11 enrolled salons.

|  | **Pre-Intervention** |  |  |  | **Post-Intervention** |  |  |  | **Ratio of GM diff (95% CI ratio GM) post-pre** |
| --- | --- | --- | --- | --- | --- | --- | --- | --- | --- |
|  | N | % below DL | AM/GM (ppm) | GSD | N | % below DL | AM/GM (ppm) | GSD |  |
| Acetone |  | 0 | 15.89/8.75 | 3.12 |  | 0 | 9.07/5.72 | 2.86 | 0.78 (0.53, 1.15) |
| MMA |  | 2 | 3.27/1.90 | 3.59 |  | 0 | 2.37/1.28 | 3.28 | 0.76 (0.47, 1.23) |
| Ethyl acetate |  | 24 | 0.35/0.25 | 2.35 |  | 30 | 0.29/0.21 | 2.41 | 0.94 (0.68, 1.30) |

## DISCUSSION

We report the process and results of our feasibility study to implement a participatory occupational health and safety intervention program tailored to the Vietnamese nail salon community in Philadelphia area. We partnered with Vietlead early in the process including their assessment of our ability to recruit 12 salons given timing of the study and short timeframe of the grant. The academic team and Vietlead co-developed the intervention program which included a two-hour in-person training for both owners and employees, a recommendation report based on salon observations conducted by the academic team, a consultation session to go over the report, and check-in calls during the three-month follow-up. We observed some increases in occupational health and safety knowledge and self-efficacy, as well as a reduction in personal exposure to some chemicals. While these results should be interpreted with caution due to our small sample size and lack of control arm, the study has achieved its primary aim of giving the academic team and community partner a valuable experience carrying out the research project together and learning to collaborate.

Our study was initially delayed to the COVID-19 pandemic because at the time the study was slated to start in September 2020, it was deemed unsafe to gather in-person by the city and state of Pennsylvania. Initial test of the training content had to be moved to Zoom platform (Huỳnh et al., 2023). We encountered significant challenges related to recruitment and thus, the time it took to recruit 12 salons was longer than initially anticipated. A majority of the enrolled salons either had a close connection to Vietlead staff or had been introduced to the program by someone they trusted. As such, involvement of trusted community-based organizations like Vietlead and individuals was critical to the success of our recruitment efforts. In addition to the reasons for refusal reported by participants, Vietlead staff also shared with the academic team that because this program was too new to the community, there was no one from the community to vet the program, which made it challenging to convince people of its potential benefits. After the participants went through the program, a few owners voluntarily spread the word about the program and told Vietlead staff of new salons who might be interested in the program in the future. While recruitment for this study was met with challenges, we are optimistic that future recruitment will be a bit easier with the help from participants in the first cohort. Vietlead trainers delivered the training in-person at the salons, usually after work hours. While this process was time-consuming, it provided Vietlead and the academic team the opportunity to build relationships with the community as relationship building is a key part of CBPR and also essential for Vietlead to work effectively within the community. At the end of study, we also organized a community appreciation event to share preliminary results and provided the academic team, community partners, and participants also connect with one another. Our future work will likely incorporate both in-person and online training to accommodate training preferences. Our research team have had experience with online training during the pandemic (Huỳnh et al., 2023). While online training was easy for most people, we have had individuals with limited technology skills and English proficiency, thus budgeting for time and effort for technology assistance (both in-person and remotely) should be part of the initial planning.

For outcome measures, we used a combination of acute health symptoms, self-reported practices and self-efficacy, and environmental changes. Given that this study has a short three-month follow-up, reductions of health symptoms are not expected to be detected as evidenced by the wide confidence intervals of the effect estimates. Rather, the acute self-reported health symptoms may help us understand the baseline health symptoms. For example, about half of the participants reported having hay fever in the past 12 months. Our prevalence rate is on par with previously reported study conducted with Asian NSW in other parts of the country. A study of Asian NSW in the Boston area reported that about 44% of their participants reported any respiratory symptoms (difficulty breather, regular cough, sinus/nasal, irritation) (Roelofs et al., 2008). Quach et al., 2008 study reported that 47% of Vietnamese nail salon workers in California reported health symptoms that may be associated with solvent exposure including skin irritations, breath problems, eye, and throat irritations. The burden of these respiratory conditions among NSW is substantially higher for wheezing, hay fever, and eczema conditions than the general population. Data from the European Respiratory Health Study that used the same instrument reported that among 193,912 adults in 43 centers in 17 countries reported the overall prevalence of symptoms at 6.6% (0.9–32.7%) for current wheeze, 4.4% (0.9–29.0%) for asthma ever, 14.4% (2.8–45.7%) for hay fever ever and 9.9% (1.6–29.5%) for eczema ever (Mortimer et al., 2022). While the prevalence for wheeze and asthma among our participants are similar to the general population, the prevalence of hay fever symptoms and eczema were approximately three times higher than the general population. It is notable that we observed a number of symptoms such as hay fever, chest tightness, eczema, and eye irritation that appear to have occupational causes due to their improvement when away from work. We may wish to target understanding these specific symptoms and implied conditions in the future.

We observed an increase in glove use but not in mask use, stretches, or going outside to get fresh out. Our high self-reported mask use (100%) at baseline is likely to due to the then on-going pandemic primarily for COVID-19 protection. As the country emerges from the pandemic and restrictions ease, participants appeared to also relax their mask use. Future education campaigns should also emphasize messaging the benefits of mask use for both chemical and/or dust and airborne pathogens protection in the salons. Our training recommended going outside to get fresh air, but we also recognize that in some cases, it may not be feasible if the salon is located in neighborhoods that may not be safe to do so or in cold weather. This stresses the importance of having good ventilation in the salon for those instances.

We observed a reduction of chemical exposure of some chemicals but our ability to attribute the reduction of exposure is extremely limited given the small sample size and lack of a control arm. There are likely confounding factors due to seasonality. For example, the baseline data collection took place mostly during winter, and when we collected follow-up data during the summer season, we did notice owners opening doors when the weather was nice. Because such behavior was not assessed at baseline, we could not ascertain how much of that was attributed to the training. In our future assessment, we will likely modify our outcome measure to help us better capture “low-hanging fruit” behaviors such as these from the owners. Our chemical exposure levels were all below the occupational exposure limits. Despite of this fact, the prevalence rates of acute respiratory and skin irritations from solvent exposures should be of concern to the community as these acute effects do impact quality of life including neurological impairment from exposure to low-level of solvents. We presented the results post-vs pre-intervention of the neurotoxicity score to estimate potential intervention effort to be consistent with the theme of this paper. A more thorough analysis is forthcoming to better understand health effects of low-level solvent exposures on the neurological system.

We conducted parallel in-depth process evaluation reported in Huynh et al., 2023. All owner participants, except one, reported to us that they did not make substantial changes to the salon environments since our initial observations. Our Vietlead trainer reported to us that the salon owner who purchased new nail salon manicure tables with built-in local exhaust ventilation (LEV) did so because the salon was already in the process of being upgraded. After learning about the function of LEV to protect health through our training, the owner decided to purchase them to protect his family who worked in the salons. Through interviews with the participants, we learned that even though the participants acknowledge the benefits of training in increasing their awareness of the hazards, there were many perceived barriers to implementing changes related to salon environment. These barriers included high cost, lack of the availability of alternative products at the local nail suppliers, lack of motivation from the owners, and work practices such as long work hours and low wage impacting employee morale and motivation for additional training (Huỳnh et al., 2023). Consideration of these barriers may require external policy to incentivize and support salons such as the voluntary Healthy Nail Salon Recognition Program (Nguyen et al., 2022). In a survey of community stakeholders in Philadelphia, we learned that all stakeholders support the creation of the program but lack of funding for the local health department to adopt and implement the program was identified as a major barrier as well as the need for the community mobilization to have their voices heard that such a program is important to the health of the immigrant communities (Nguyen et al., 2022).

Our study has several limitations. First, the salons and participants in the study may be early adopters and may be not representative of salons in the region. Second, self-reported measures of health symptoms, behaviors, and self-efficacy may suffer from recall bias. We cannot exclude correlated errors in questionnaire responses, such that persons under or over-reporting at one point in time, exhibiting the same tendency at another time. While we used validated research instruments whenever possible, there is lack of evidence of validity for the Vietnamese population, although this concern is mitigated by the fact that some instruments have a history of successful use in diverse populations.

Despite these limitations and being a small sample, our study’s participants reflected the demographic characteristics of the overall nail salon workforce. Most of them are foreign-born women with limited English proficiency and low socioeconomic status. The vulnerability of this workforce calls for a comprehensive and flexible approach to workplace and public health interventions. Our community-based participatory research approach, which prioritizes building trusting relationships with their community partners, understanding the significance of shared experiences, and taking into accounts fears and stigma of being immigrant workers, allowed the participants to be empowered and included in the decision-making process. This allowed us to better understand and work closely with the community, but also required us to be flexible and adjust our study design to meet their needs (Katigbak et al., 2016; Vaughn et al., 2017). Future research funding opportunities should consider the benefits of community-based participatory research and encourage researchers to explore innovative ways to ensure their studies are truly inclusive, equitable and relevant to the communities they address.

## CONCLUSION

Our research study demonstrates a participatory approach to designing and implementing an occupational health intervention study for the underserved nail salon worker population in the Philadelphia area. Getting buy-in from community leaders and community partners early in the process was a key factor in our recruitment success. Our outcome assessments, while small in sample size, provide early evidence of potential effectiveness of the intervention for behavioral outcomes. Future use of intervention materials will likely need further tailoring but overall, the multi-level intervention focusing offering support to owners and community organizations offer a promising approach to work with primarily immigrant small businesses owners and their employees.

## Data Availability

All data produced in the present study may be available upon reasonable request to the authors.

## Acknowledgements

We are grateful for the continued support and guidance from Vietlead staff (Tracy Nguyen, Van Long, Ngoc Nguyen), and students and volunteers (Raveena Suman, Chau Nguyen, Mai Anh Nguyen, Hoang Anh Nguyen), advisory board members, salon owners and workers in the study, as well as those from the nail salon community in the Greater Philadelphia region. We would also like to thank the following individuals and organizations who have not only shared their research instruments, technical resources, and expertise but also have been the inspiration for us: Dr. Thu Quach at the Asian Health Services, Mr. Tuan Nguyen, the California Healthy Nail Salon Collaborative, and Mrs. Trisha Le from the National Nail Association of America.

## Funding

Our study was generously funded by the U.S. National Institute of Occupational Safety and Health through the grants K01OH011191 and R21OH011740, Drexel University Equipment Grant, and Dr. Arthur Frank, the Drexel University Urban Health Collaborative Pilot Study. This paper is the sole view of the authors and does not reflect the views of the funders.

## Notes

### Competing Interest Statement

The authors have declared no competing interest.

### Author Declarations

Ethics committee/IRB of Drexel University gave ethical approval for this work.

